# High-pass filter settings and possible mechanism of discrete electrograms in left bundle branch pacing

**DOI:** 10.1101/2022.09.01.22279483

**Authors:** Jiabo Shen, Longfu Jiang, Hao Wu, Hengdong Li, Lu Zhang, Jinyan Zhong, Shanshan Zhuo, Lifang Pan

## Abstract

**Objective:** The characteristics of discrete intracardiac electrograms in selective left bundle branch (SLBB) pacing (SLBBP) have not been described in detail previously. This study aimed to examine the effect of different high-pass filter (HPF) settings on discrete ventricular components in an intracardiac electrogram (EGM) and to analyze its possible mechanisms.

**Methods:** This study included 95 patients with indications of permanent cardiac pacing. EGMs were collected at four different HPF settings (30, 60, 100, and 200 Hz) with a low-pass filter at 500 Hz, and their possible mechanisms were analyzed.

**Results:** LBBP was successfully achieved in 92.6% (88/95) of patients. SLBBP was achieved in 80 patients. The occurrence rates of discrete EGM were 18.9%, 40.0%, 74.7%, and 84.2% for HPF settings of 30 Hz, 60 Hz, 100 Hz, and 200 Hz, respectively. The analysis of discrete ECG detection showed significant differences between the different HPF settings. By using the discrete EGM as the SLBB capture golden standard, the results of EGMs revealed that the 30 Hz HPF has a sensitivity of 23% and specificity of 100%. The 60 Hz HPF had a sensitivity of 48% and a specificity of 100%. The 100 Hz HPF had a sensitivity of 89% and a specificity of 100%. The 200 Hz HPF had a sensitivity of 100% and specificity of 100%.

**Conclusions:** An optimal HPF setting of 200 Hz is recommended for discrete electrogram detection. A discrete EGM should exhibit an isoelectric interval. A **steep** deflection and spinous ventricular EGM morphology nearly identify an intrinsic EGM morphology.

## Introduction

Left bundle branch (LBB) pacing (LBBP) is a novel native conduction system pacing strategy.[1] Changes in the intracardiac electrograms (EGMs) are usually assessed during LBBP implantation via an electrophysiological recording system (EPS) to confirm selective LBB capture or monitor the current of injury (COI).[2, 3] A discrete EGM and an isoelectric interval have been previously used as criteria to confirm the selective LBB (SLBB) capture, which suggests that only the conduction system was captured, and the myocardium was lost. Therefore, identifying discrete EGM is crucial for accurately diagnosing SLBB capture. [4] Filtered unipolar electrograms were obtained in previous studies, usually with settings of 30 and 100/300/500 Hz [5-7], for LBBP. However, filtering could remarkably change the morphology of the ventricular EGM, introducing the possibility of errors in the evaluation of electrograms. When the high-pass filter (HPF) of the LBB lead channel is set to 30 Hz and clipping is set at 3 cm, the entire ventricular endocardial signal may not be observed owing to the large amplitude in the EGM. Therefore, identifying discrete EGMs is a challenging task. We hypothesized that an HPF other than 30 Hz improved the detection of discrete EGM. Therefore, in the present study, we aimed to evaluate the accuracy of discrete EGM in diagnosing selective LBBP (SLBBP) in different HPF settings and analyze its possible mechanisms.

## Methods

### Patient population and definition of LBB capture

This retrospective observational study enrolled consecutive patients who underwent successful permanent pacemaker implantation. LBBP uses the continuous pacing and recording technique, and the procedure has been described by us in detail elsewhere.[8-10] The study protocol was approved by the Ethics Committee of the Hwa Mei Hospital, Ningbo, China. Written informed consent was obtained from all patients.

Successful LBBP is defined as follows. LBB capture is characterized by paced QRS morphology of the RBBB pattern and all of the following criteria: (1) differential pacing at 8 and 2 V, producing the shortest and constant V6 R-wave peak time (preferably <75 ms), and (2) demonstration of left ventricular septal (LVS) to non-selective LBB capture transition during constant output and non-selective to selective LBB capture during unipolar pacing threshold assessment. [5, 11]

### Data recording and analysis

The baseline patient characteristics and indications for pacing were documented. Twelve-lead electrocardiogram (ECG) and EGM from the pacing lead were continuously recorded with an EPS (EP-Workmate, Abbott Laboratories, Chicago, IL, USA). For each patient, the differences in discrete EGM morphologies were collected and analyzed using four different HPF filters (30 Hz, 60 Hz, 100 Hz, and 200 Hz), and the low-pass filter was set at 500 Hz. To ensure high precision, the analysis of discrete EGM morphology was performed using endocardial channel recording, digital calipers, fast sweep speed (200 mm/s), and appropriate signal augmentation. The clipping was set at 3 cm, and the amplitude was set at 0.5 mV/cm.

The characteristics of the various transitions in discrete EGM morphology were analyzed retrospectively after the procedure. All EGM morphologies were independently analyzed by two medical practitioners who were highly experienced in EGM interpretation. When (1) the isoelectric interval, (2) the paced initial steep deflection, and (3) spinous ventricular EGM nearly identical to the intrinsic ventricular EGM could be observed independently by both doctors at different HPF filters, the observation was marked as a discrete EGM (patients with LBBB meeting criteria 1 and 2 because intrinsic LBB conduction cannot be observed). The EGM readers were blinded to the study’s purpose. In the absence of concordance between the two readers, a third cardiologist practitioner adjudicated the results.

### Statistical analysis

All continuous data are presented as mean ± standard deviation (SD). Categorical data were presented as numbers and percentages. We used Student’s t-test to compare continuous variables. To evaluate the diagnostic accuracy of detecting discrete EGM, the sensitivity, specificity, positive predictive value (PPV), and negative predictive value (NPV) of different HPF were calculated. A *p*-value < 0.05 was considered statistically significant. The statistical software IBM SPSS Statistics for Windows (version 26.0, IBM Corp, Armonk, NY, USA) was used for analysis.

## Results

From April 2021 to March 2022, data for 95 patients who underwent pacemaker implantation were consecutively and retrospectively collected from a single institution (Hwa Mei Hospital, University of Chinese Academy of Sciences). Their mean age was 74.2 ± 8.6 years, and 39/95 (41.1%) were females. Clinical and procedure-related characteristics of the study population are shown in **Table 1**. Successful LBBP with evidence of LBB system capture was achieved in 88 patients (92.6 %). SLBBP was achieved in 80 patients during the threshold testing. Eight patients were confirmed as having NSLBBP. In seven patients, LBBP failed because of the inability to capture the LBB. These patients eventually underwent LVS pacing. The pacing indications in the 88 patients who achieved LBBP were sick sinus syndrome in 25 (28.4%), atrioventricular block in 53 (60.2%), atrial fibrillation with bradycardia in 7 (8.0%), and heart failure in 3 (3.4%). LBB potential (Po_LBB_) was recorded in 65 (73.9%) of 88 patients.

The performance of the discrete EGM detection with different HPF is shown in **Figures 3-6**. The discrete EGM occurrence rates for different EGM setup channels were compared. The results of the discrete EGM detection are summarized in **Tables 2 and 3**. The occurrence rates of discrete EGM were 18.9% (18/95), 40.0% (38/95), 74.7% (71/95), and 84.2% (80/95) for HPF settings of 30 Hz, 60 Hz, 100 Hz, and 200 Hz, respectively (**Table 2**). The analysis of discrete ECG detection showed significant differences between the different HPF settings (**Table 3**). Using the discrete EGM as the gold standard, the results of EGMs indicated that the 30 Hz HPF had a sensitivity of 23% and specificity of 100%. Furthermore, the PPV was 100% and 19%, respectively. The 60 Hz HPF had a sensitivity of 48% and specificity of 100% for SLBB capture, with a PPV of 100% and NPV of 26%. The 100 Hz HPF had a sensitivity of 89% and specificity of 100%, with a PPV of 100% and NPV of 63%. The 200 Hz HPF had a sensitivity of 100% and specificity of 100% for SLBB capture, with a PPV of 100% and NPV of 100%.

**Figure 1.**
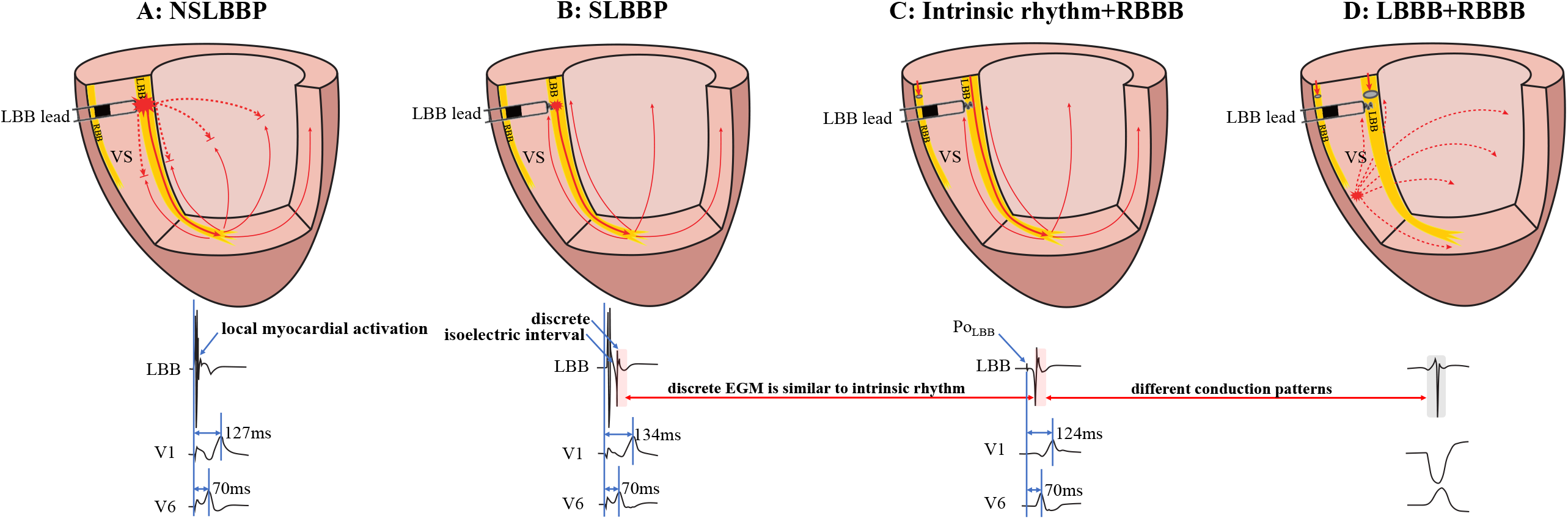
Schematic diagram of different conduction patterns in a patient with RBBB and intermittent LBBB. A: NSLBBP captures both the local myocardium and LBB without the presence of isoelectric interval and discrete EGM. B: In SLBBP, pacing electrical stimulation is conducted through the conduction system to the apex and propagates to the base of the interventricular septum. Isoelectric interval and discrete EGM were observed. C: Intrinsic conduction antegrade to the apex and propagates to the base. Morphology of intrinsic and paced ventricular EGMs was nearly identical (red rectangle). D: The EGM morphology of LBBP is similar to the native rhythm but different from the EGM morphology of LBBB (red and grey rectangle). NSLBBP: non-selective left bundle branch pacing; SLBBP: selective left bundle branch pacing; EGM: intracardiac electrogram; LBB: left bundle branch; LBBB: left bundle branch block; RBBB: right bundle branch block; VS: ventricular septum.

**Figure 2.**
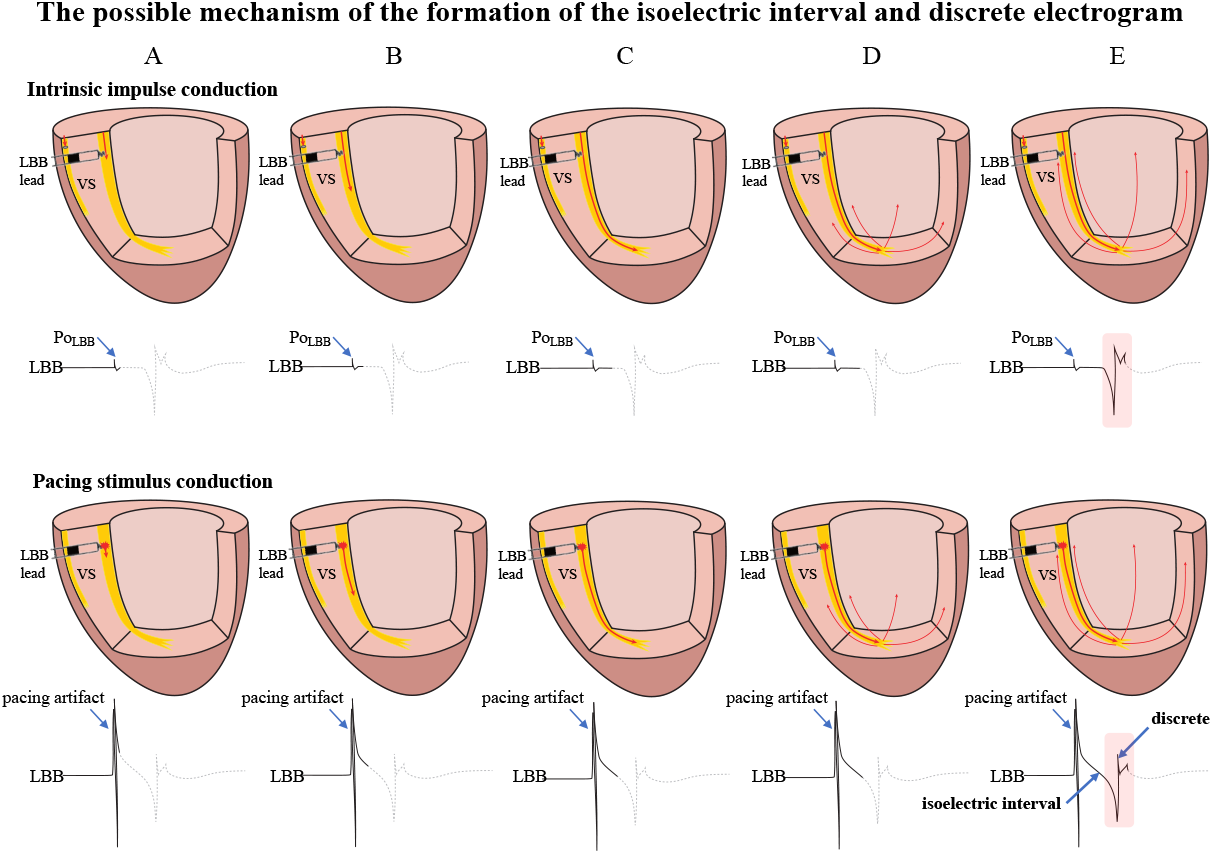
The possible mechanism of the formation of the isoelectric interval and discrete electrogram. A: Intrinsic impulse or pacing stimulus in the LBB is sensed by the tip lead. B-D. The formation process of the isoelectric interval. E: Intrinsic impulse or pacing stimulus reaches the distal end of the His-Purkinje system, excites the apical myocardium, propagates into the basal myocardium, and is then sensed by the tip lead, manifesting as an isoelectric interval and discrete electrogram. Abbreviations as in Figure 1.

**Figure 3.**
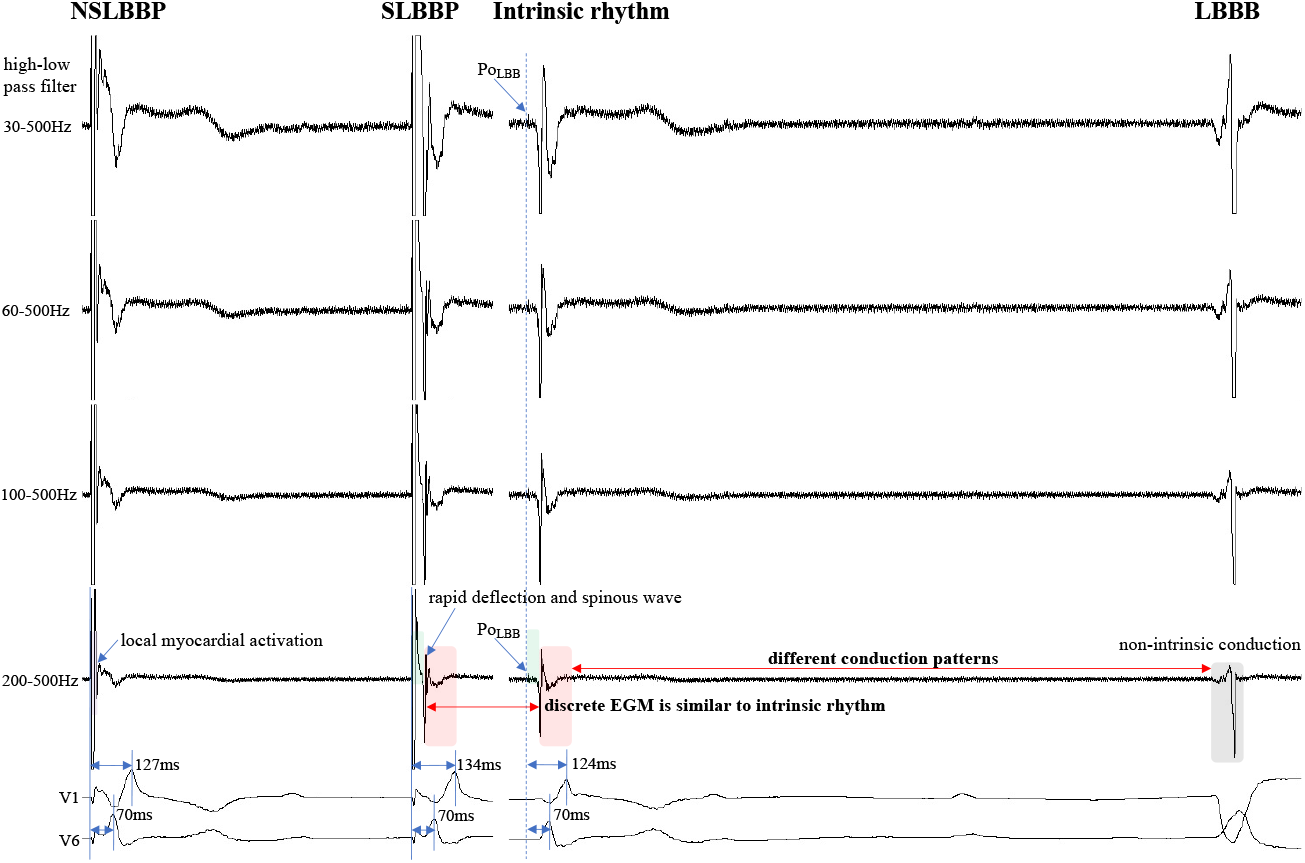
Discrete EGM morphology resembles ventricular EGM morphology of intrinsic rhythm (red rectangle). The EGM morphology of LBBP is similar to the native rhythm but is completely different from the EGM morphology of LBBB (red and grey rectangle). Po_LBB_ can be observed at different high-pass filter settings (blue dashed line). Isoelectric interval does not appear during local myocardial activation (purple rectangle). Isoelectric interval is affected by pacing artifacts (green rectangle). Abbreviations as in Figure 1.

## Discussion

LBBP includes non-selective LBBP (NSLBBP) and SLBBP. In NSLBBP, the local ventricular myocardium is directly captured by the pacing stimulus and is not seen as a discrete component in the EGM because of the surrounding ventricular tissue capture (**Figure 1A**). SLBBP was defined as only capturing the LBB with a typical RBBB morphology as well as a discrete component between the pacing stimulus and the onset of local ventricular activation due to local myocardium not being directly captured in the EGMs (**Figure 1B**). A discrete EGM is a characteristic of an SLBBP [2]. It appears as a current deflection wave with a short isoelectric interval and large amplitude with an HPF setting of 30 Hz.

The isoelectric interval is often observed between the pacing artifact and the paced discrete ventricular component. This phenomenon includes the true isoelectric interval (time required for immediate peri-electrode tissue excitation or a local response) and local conduction time (time required for propagated excitation to recruit sufficient local myocardial tissue to produce the ventricular EGM).[12] Typically, the isoelectric interval is short (<30 ms).[5] An increased isoelectric interval may result from nonhomogeneous excitation propagation from the stimulation site and conduction delay in the His–Purkinje system.[12] A previous study positioned a linear multielectrode catheter along the left ventricular septum to record intracardiac signals from the base to the apex to assess left ventricular activation sequences.[13] According to the “V”-shaped conduction pattern observed in this study, the mechanism underlying the formation of the isoelectric interval may be associated with the propagation of impulse or pacing stimulus through the conduction system, reaching the distal part of the His–Purkinje system to excite the apical myocardium, and subsequent propagation to the basal myocardium in the interventricular septum by the electrode sensing (**Figure 2**). In non-LBBB patients, discrete ventricular EGM was nearly identical to the native ventricular EGM morphology (**Figure 1-6, red rectangle**). Intrinsic and paced ventricular EGMs were nearly identical in patients with intermittent LBBB with native anterograde conduction (**Figure 3, red rectangle**). Therefore, we speculate that in patients with complete LBBB, although the native rhythm (intrinsic conduction) cannot be observed, the EGM morphology of the intrinsic LBB conduction should be consistent with the EGM morphology of LBB pacing. Additionally, we observed inconsistent ventricular EGM morphology on the tip lead due to different LBBB and intrinsic LBB conduction pathways (**Figure 3, gray rectangle**).

In previous studies, high- and low-pass filter settings of 30 Hz and 500 Hz were set to record the EGM. Clinicians usually employ a 30 Hz HPF to record Po_LBB_ .[6] However, with this HPF, discrete EGMs may be missed easily because the clipping level limits the display range and does not allow the user to view large-amplitude endocardial signals (**Figures 4 and 5, blue rectangle**). Additionally, in some patients, demonstration of the isoelectric interval and discrete local electrogram may be challenging due to short stimulus to ventricular intervals, effects of stimulus artifact, and far-field recording by the pacing lead (**Figure 3-5, green rectangle**).[14] In contrast, pacing artifacts and separation of ventricular components can be observed in some cases, which are easily confused with true discrete EGM (**Figures 4 and 5, red dashed frame**). However, these observations do not represent the SLBB capture. Based on the observations in our study, the paced initial steeply deflected ventricular EGM morphology should be nearly identical to the intrinsic ventricular EGM morphology with an isoelectric interval to be considered SLBBP (**Figures 1-6, red rectangle**).

**Figure 4.**
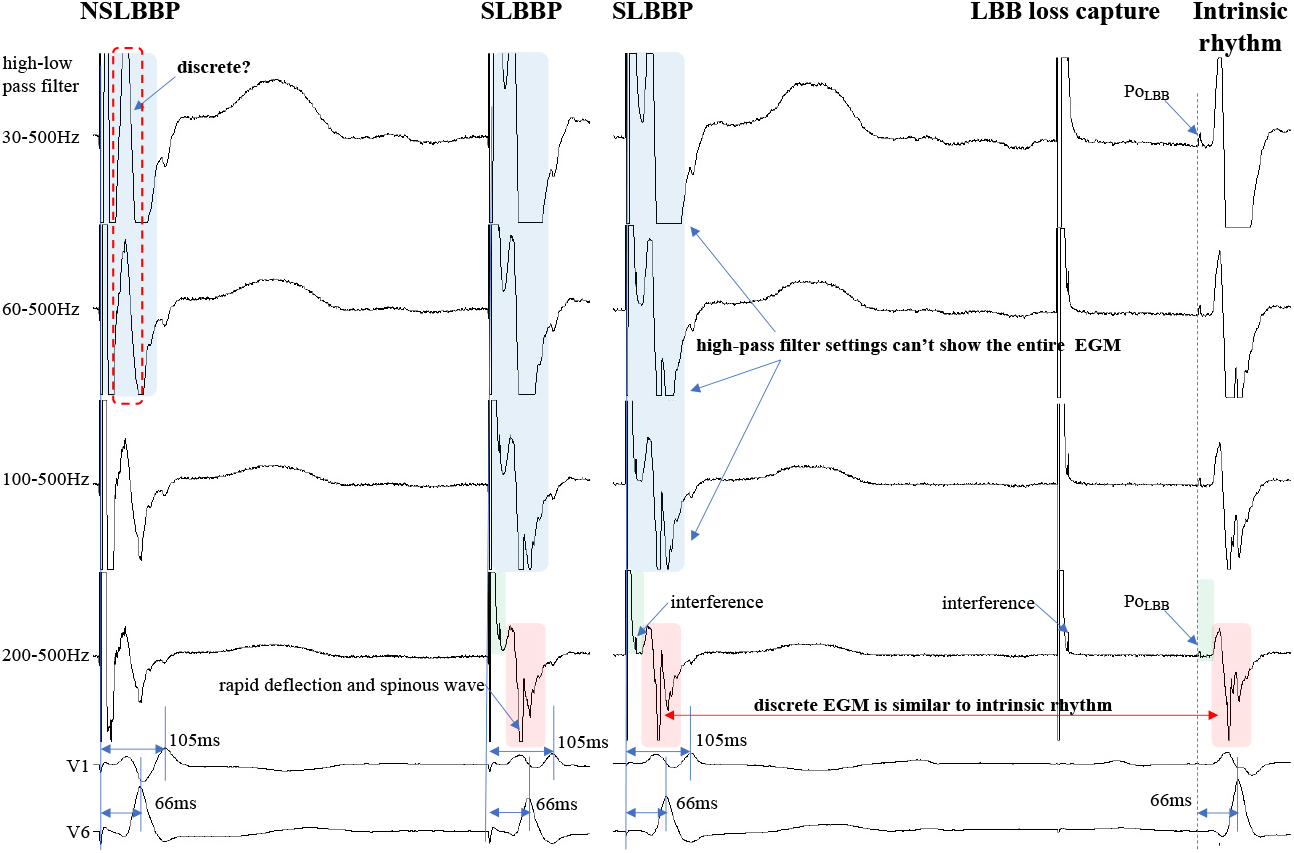
Discrete ventricular components cannot be accurately identified because the entire EGM morphology cannot be displayed (blue rectangle). **Steep** deflection and spinous discrete EGM morphology are similar to ventricular EGM morphology of intrinsic rhythm (red rectangle). Isoelectric interval is affected by pacing artifacts (green rectangle). Po_LBB_ can be observed at different high-pass filter settings (blue dashed line). Abbreviations as in Figure 1.

**Figure 5.**
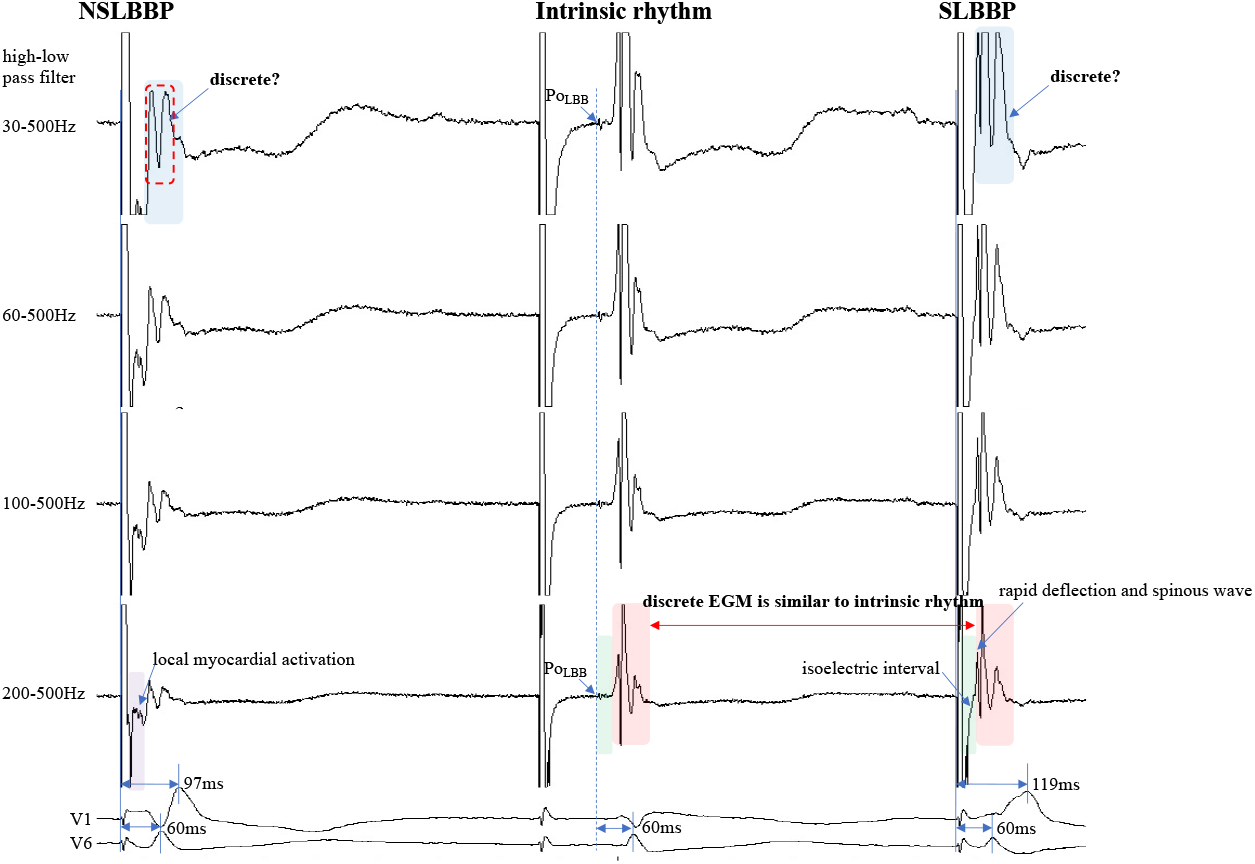
Discrete ventricular components cannot be accurately identified because the entire EGM morphology cannot be visualized (blue rectangle). Discrete EGM morphology resembles ventricular EGM morphology of intrinsic rhythm (red rectangle). The isoelectric interval of paced rhythm and native rhythm (green rectangle). Changes in high-pass filter settings do not affect the identification of Po_LBB_ (blue dashed line). Isoelectric interval does not appear during local myocardial activation (purple rectangle). Abbreviations as in Figure 1.

**Figure 6.**
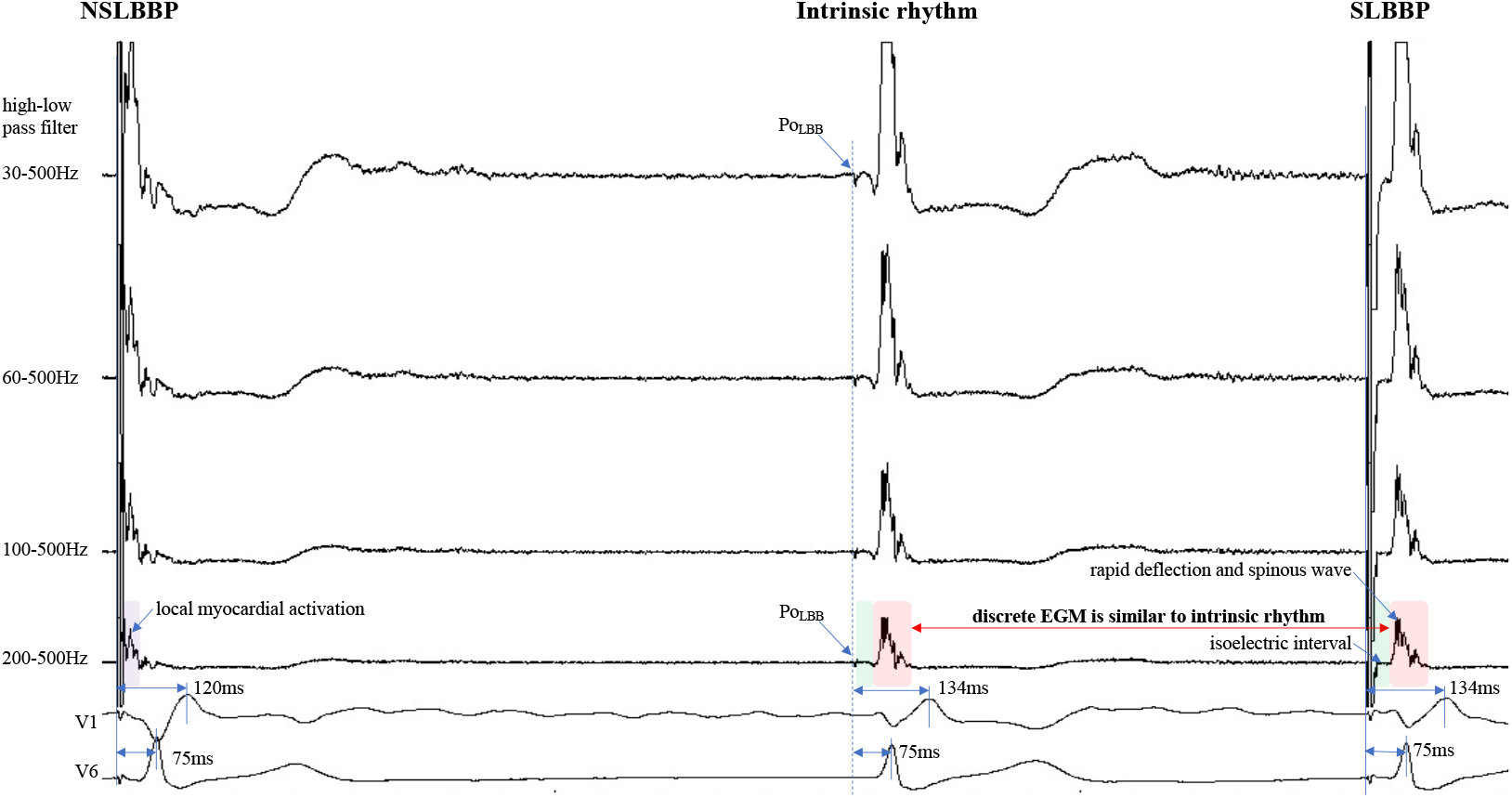
The discrete EGM morphology is nearly identical to the ventricular EGM morphology of the intrinsic rhythm (red rectangle). Isoelectric intervals of paced rhythm and intrinsic rhythm are nearly identical (green rectangle). Isoelectric interval does not appear during local myocardial activation (purple rectangle). Po_LBB_ can be observed at different high-pass filter settings (blue dashed line). Abbreviations as in Figure 1.

The HPF is designed to eliminate unwanted lower frequencies by allowing frequencies higher than the filter settings to pass. The higher the frequency, the lower the baseline wander. The low-pass filter passes frequencies lower than the cutoff frequency and attenuates higher frequencies. The lower the frequency, the lower is the baseline noise. The change in the signal produced by filtering depends on the frequency of the unfiltered signal. Variations in the HPF produced marked changes in electrogram morphology, introducing the possibility of inaccurately assessing discrete EGM (**Figures 4 and 5, blue rectangle**). Accurate interpretation of discrete EGMs highly depends on the magnitude of the ventricular component. An excessive amplitude affects the identification of a discrete EGM. Therefore, with clipping set to 3 cm and amplitude set to 0.5 mV/cm, we attempted to show the intact and entire ventricular EGM more clearly by adjusting the HPF setting to confirm SLBB capture by identifying discrete EGM and isoelectric interval.

To the best of our knowledge, no previous study has identified a discrete EGM by adjusting the band-pass filter with different setup conditions in LBBP. One purpose of this study was to define the impact of different HPF settings on the accuracy of discrete electrogram identification. These parameters were compared for different HPF values of 30, 60, 100, and 200 Hz. The unipolar electrogram signal morphology was subsequently analyzed. Our research suggested that the occurrence rates of discrete EGM were 18%, 40%, 71%, and 84% for HPF settings of 30 Hz, 60 Hz, 100 Hz, and 200 Hz, respectively. A low rate of discrete EGM identification at 30 Hz HPF was observed (sensitivity, 23%; specificity, 100%). However, the 200 Hz HPF had a sensitivity of 100% and specificity of 100% for SLBB capture. The result suggest that a 200-Hz filter may be a desirable choice. Moreover, a discrete Purkinje potebtial precedes the onset of local ventricular electrography. Filtered unipolar electrograms were obtained at 30 Hz and 500 Hz to record the Po_LBB_ .[6] A low-pass filter was used to eliminate the noise. In our study, the low-pass setting was 500 Hz, although the HPF settings differed. This suggests that such a setting does not affect the identification of the Po_LBB_ (**Figure 3-6, red, blue dashed line**). Decreasing the signal gain and lowering the low-pass filter setting makes the discrete EGM more visible, but has the disadvantage of not being able to observe the Po_LBB_ accurately.

## Study limitations

This study has several limitations. This retrospective study was performed at a single center and included a relatively small number of patients. Although the intrinsic and paced EGMs were also nearly identical in patients with intermittent LBBB block, there is still a lack of evidence suggesting that paced discrete EGM is nearly identified as native EGM in patients with complete LBBB. We used only one particular manufacturer EPS. It is possible that the ability to detect discrete EGM might differ depending on the EPS used because of differences in signal processing algorithms. Therefore, it is unknown whether the results of this study can be extended to other patient groups or different EPSs. To address these issues, a larger study including different patient groups and EPSs is needed. Randomized controlled and prospective trials are needed to confirm the findings of this study and to provide guidance to clinicians. Outcomes data in terms of persistence of the sensed findings, ventricular function, or quality of life-based are lacking.

## Conclusion

We demonstrated that an HPF setting of 30 Hz, routinely used in clinical practice, cannot reliably meet the clinical requirements of discrete EGM detection. Our results suggest that clinicians can adjust HPF appropriately to improve discrete EGM diagnosis, and a 200-Hz filter may be a desirable choice. A discrete EGM should show an isoelectric interval, and a steep deflection and spinous ventricular EGM morphology nearly identify an intrinsic EGM morphology.

## Data Availability

All data produced in the present study are available upon reasonable request to the authors

