## Supplementary material for "High-pass filter settings and possible mechanism of discrete electrograms in left bundle branch pacing": 1

| **Table 1.** Baseline characteristics, pacing indications, and baseline echocardiography and ECG data of patients who underwent attempts at LBBP. Patients who underwent LVSP were those in whom LBBP failed. | | LBBP  (n = 88) | | LVSP  (n = 7) | | *p* | |
| --- | --- | --- | --- | --- | --- | --- | --- |
| Age (years) | | 74.0 ± 8.5 | | 77.3 ± 10.6 | | 0.39 | |
| Male | | 53 (60.2) | | 3(42.9) | |  | |
| Pacing indication (n) | |  | |  | |  | |
| Atrioventricular block | | 53 (60.2) | | 6(85.7) | |  | |
| Sick sinus syndrome | | 25 (28.4) | | 0(0) | |  | |
| Atrial fibrillation with bradycardia | | 7 (8.0) | | 1(14.3) | |  | |
| Heart failure | | 3 (3.4) | | 0(0) | |  | |
| Comorbidities (n) | |  | |  | |  | |
| Hypertension | | 53 (60.2) | | 6(85.7) | |  | |
| Diabetes mellitus | | 21 (23.8) | | 1(14.3) | |  | |
| Cardiomyopathy | | 5 (5.7) | | 0(0) | |  | |
| Coronary heart disease | | 14 (15.9) | | 1(14.3) | |  | |
| Atrial fibrillation | | 28 (31.8) | | 2(28.6) | |  | |
| LVEF (%) | | 63.7 ±10.2 | | 66.6 ± 5.5 | | 0.35 | |
| LVDD (mm) | | 50.2 ± 7.7 | | 50.3 ± 4.0 | | 0.13 | |
| QRS morphology (n) | |  | |  | |  | |
| Narrow QRS | | 60 (68.2) | | 6(85.7) | |  | |
| RBBB | | 21 (23.9) | | 0(0) | |  | |
| LBBB | | 14 (15.9) | | 0(0) | |  | |
| NIVCD | | 2 (2.3) | | 1(14.3) | |  | |
| Procedure-related parameters | |  | |  | |  | |
| LBB potential observed (n) | | 65 (73.9%) | | 0(0) | |  | |
| Threshold (V/0.5ms) | | 0.58 ± 0.25 | | 0.57 ± 0.20 | | 0.58 | |
| R-wave amplitude (mV) | | 14.7 ± 6.8 | | 9.6 ± 4.0 | | 0.12 | |
| Impedance (Ω) | | 723.0 ± 127.7 | | 759.6 ± 128.3 | | 0.93 | |
| Lead depth (mm) | | 14.7 ± 2.7 | | 14.7±1.5 | | 0.08 | |
| LVEF, left ventricular ejection fraction; LVDD, left ventricular end-diastolic dimension; RBBB, right bundle branch block; LBBB, left bundle branch block; NIVCD, non-specific intraventricular conduction disturbance; LBB, left bundle branch; LBBP, left bundle branch pacing; LVSP, left ventricular septal pacing. Continuous data were presented as mean ± standard deviation. *p* < 0.05 was considered statistically significant. | | | | | | | |
| **Table 2.** Detection of discrete EGM at different high-pass filter settings. | | | | | | | |
|  | 30Hz | | 60Hz | | 100Hz | | 200Hz |
| Presence of discrete EGM | 18 (18.9) | | 38 (40.0) | | 71 (74.7) | | 80 (84.2) |
| Absence of discrete EGM | 77 (81.1) | | 57 (60.0) | | 24 (26.3) | | 15 (15.8) |
| Data are presented as numbers (%). EGM: intracardiac electrogram. | | | | | | | |

| **Table 3.** Results and diagnostic accuracy of different high-pass filter for detecting discrete EGM. | | | | |
| --- | --- | --- | --- | --- |
|  | 30Hz | 60Hz | 100Hz | 200Hz |
| Sensitivity % (95% CI) | 0.23 (0.14-0.33) | 0.48 (0.36-0.59) | 0.89 (0.79-0.94) | 1.00 (0.94-1.00) |
| Specificity % (95% CI) | 1.00 (0.75-1.00) | 1.00 (0.75-1.00) | 1.00 (0.75-1.00) | 1.00 (0.75-1.00) |
| PPV % (95% CI) | 1.00 (0.78-1.00) | 1.00 (0.89-1.00) | 1.00 (0.94-1.00) | 1.00 (0.94-1.00) |
| NPV % (95% CI) | 0.19 (0.12-0.30) | 0.26 (0.16-0.40) | 0.63 (0.41-0.80) | 1.00 (0.75-1.00) |
| 95% CI: 95% Confidence interval; EGM: intracardiac electrogram; PPV: Positive predictive value; NPV: Negative predictive value. | | | | |
